# Genomic Diversity of the SARS-CoV-2 in Turkey and the Impact of Virus Genome Mutations on Clinical Outcomes

**DOI:** 10.1101/2020.12.25.20248851

**Authors:** Ilker Karacan, Tugba Kizilboga Akgun, Nihat Bugra Agaoglu, Payam Zolfagharian, Mehtap Aydin, Gizem Alkurt, Jale Yildiz, Betsi Kose, Nisan Denizce Can, Ayse Serra Ozel, Nilsun Altunal, Arzu Irvem, Yasemin Kendir Demirkol, Ozlem Akgun Dogan, Levent Doganay, Gizem Dinler Doganay

## Abstract

COVID-19 is a viral respiratory disease caused by SARS-CoV-2 infection. Global efforts of genomic surveillance of the virus give chance to track the spread of the pandemic. Global emergence of some viral mutations called attention and various studies have been suggested about increased infectivity of the virus. Herein, we sequenced viral genomes isolated from 184 patients in Istanbul and analyzed clinical metadata for the investigation of any viral mutation which affects the disease course of the host. We did not detect any viral mutations affecting the disease outcome in our cohort. Besides, we observed intra-host mutations in 76% of the isolates. Insertion/deletion and stop-gain mutations are also significantly less common among intra-host variants compared to consensus viral genome mutations. Longitudinal genomic surveillance is essential for timely detection of any lineages that might affect clinical outcome, the performance of diagnostic assays, or even the immunological escape of the virus.

## Introduction

Since the first case at the end of 2019, the novel betacoronavirus SARS-CoV-2 has spread the world rapidly by reaching more than 60 million affected people and 1.4 million deaths as of November 2020. Host response to SARS-CoV-2 infection and clinical severity can show high heterogeneity. While many cases do not show any symptoms, some COVID-19 patients develop acute respiratory distress syndrome and other life-threatening symptoms such as pulmonary edema or thrombotic coagulopathy (Huang et al., 2020). Older age, cardiovascular disease, respiratory diseases, diabetes are among known risk factors for COVID-19 clinical course (Jordan et al., 2020; Wu and McGoogan, 2020). Pathogenesis of the COVID-19 is still poorly understood but older age, male sex, and several other factors have been shown associated with increased mortality rates (Li, X. et al., 2020; Zhou et al., 2020). Evidence on multisystemic involvement of the disease provoked questions on the association between host genetic factors with clinical outcome (Initiative, 2020; Zeberg and Paabo, 2020). Host genetic factors and their pathogenesis in disease severity is still needed to be elucidated.

It has been known that RNA viruses have a much higher mutation rate, which can provide enhanced adaptation and virulence, compared to DNA viruses (Duffy, 2018; Lauring and Andino, 2010). Mutations in the RNA-based genome of coronaviruses can arise due to replication errors which may be reduced in coronaviruses due to their polymerases with proofreading mechanism (Minskaia et al., 2006; Snijder et al., 2003). Also, the RNA editing mechanism, which is a part of the host immunity, can trigger new mutation events in the viral genome (Harris and Dudley, 2015; Mangeat et al., 2003). Global efforts for sequencing SARS-CoV-2 genomes revealed that may be expected for a coronavirus, genome diversity is very low (Fauver et al., 2020). However, excessive global spreading can give the virus a chance for a natural selection of advantageous mutations resulting in increased infectivity.

High throughput sequencing methods have been progressively used during the COVID-19 outbreak. Extensive viral genome analyses by laboratories all over the world have reached the identification of more than 225,000 records in the GISAID SARS-CoV-2 database in just one year. In the meantime, several questions about viral mutations and their effects on the patient’s clinical outcomes have arisen. Recent studies claimed increased infectivity or fatality rates caused by a viral p.D614G mutation of spike protein (Becerra-Flores and Cardozo, 2020; Korber et al., 2020; Li, Q. et al., 2020). The p.D614G mutation, which is now dominating the global pandemic, has shown that it was present in retrospectively-sampled isolates in China in late January (Volz et al., 2020). However, there is not any consensus and solid evidence on the effects of this mutation in terms of infectivity or mortality rates among patients due to uncertainty whether the frequency of this mutation is caused by chance or natural selection (Grubaugh et al., 2020).

A global and continuous effort is needed for viral strain monitoring to detect any mutation which may lead to higher mortality or infectivity rate. In this study, we sequenced 184 viral genomes sampled mostly in Istanbul, between the 13th of April and the 24th of June 2020. All related clinical data has been collected and viral genome mutations were tested for any association with the patient’s clinical outcome.

## Methods

### Patients and clinical evaluation

All patients enrolled in the study evaluated with the criteria defined by ‘COVID-19 Diagnosis and Treatment Guide’ published by the Turkish Ministry of Health. We grouped the patients as asymptomatic, mild, or severe according to the following conditions:

1. Asymptomatic: Patients without any clinical, radiological symptoms or any signs of the disease.
2. Mild: Patients with mild symptoms had respiratory symptoms such as cough and dyspnea, fever, headache, sore throat, runny nose, muscle and joint pain, extreme weakness, alteration in sense of smell and taste, diarrhea.
3. Severe: Patients with severe symptoms had (1) respiratory rate ≥ 30 breaths/min; (2) SpO2 ≤ 93% while breathing room air; (3) PaO2/FiO2 ≤ 300 mmHg; (4) respiratory failure which requires mechanical ventilation; (5) shock; (6) combined with other organ failures, need to be admitted to ICU.

### Viral RNA extraction, genome sequencing, and data analysis

Viral RNA extraction, library preparation, and sequencing were performed as described previously (Karacan et al., 2020). Briefly, RNA was extracted from nasopharyngeal swab samples using the High Pure Viral RNA Kit (Roche Life Science). The sequencing library was prepared with multiplexed PCR based target enrichment using CleanPlex SARS-CoV-2 Library Preparation Kit (Paragon Genomics Inc). Pooled libraries were sequenced on the Illumina NextSeq500 instrument.

Raw sequencing data were first demultiplexed and quality assessment was performed using the FASTQC program (Andrews, 2010). Adapter sequences were trimmed using cutadapt (Martin, 2011) and reads were aligned to the reference SARS-CoV-2 genome (NC_045512.2) using bwa-mem (Li, 2013). Primers were pruned using TrimPrimers script within the fgbio tool (http://fulcrumgenomics.github.io/fgbio/). Variant calling and consensus sequence generation were performed using Samtools (Li et al., 2009) and iVAR (Grubaugh et al., 2019). Alternate bases with 10% frequency were called as variants and consensus sequences were generated for major alleles with at least 10x depth of coverage. Variant files for each isolate were converted to vcf files, which was further processed with BCFtools (Li, 2011) and annotated using ANNOVAR (Wang et al., 2010). Consensus genome sequences of isolates were aligned using MAFFT (Katoh et al., 2019). The neighbor[joining method was used to estimate phylogenies using aligned sequences and the tree was visualized using FigTree v1.4.4. Clade annotations were performed using Nextclade tool (https://clades.nextstrain.org).

### Statistical analysis

Descriptive statistics were used to present clinical data. Proportions and percentages were used for the categorical data and mean values and standard deviations were used for the continuous data. Statistical significance of categorical data was calculated by two-tailed Fisher’s exact test. A value of p < 0.05 was considered statistically significant.

## Results

Samples were collected from 184 patients in Turkey between the 13th of April and the 24th of June 2020. The majority of the samples were collected from Istanbul, the epicenter of the pandemic in Turkey. Overall, the study cohort includes 14 patients (8%) who did not manifest any symptoms, 65 patients (%37.6) have at least one comorbidity. Most of the patients (n=128, 73.1%) have a mild disease course. Detailed clinical features of enrolled patients were presented in Table 1.

**Table 1:**
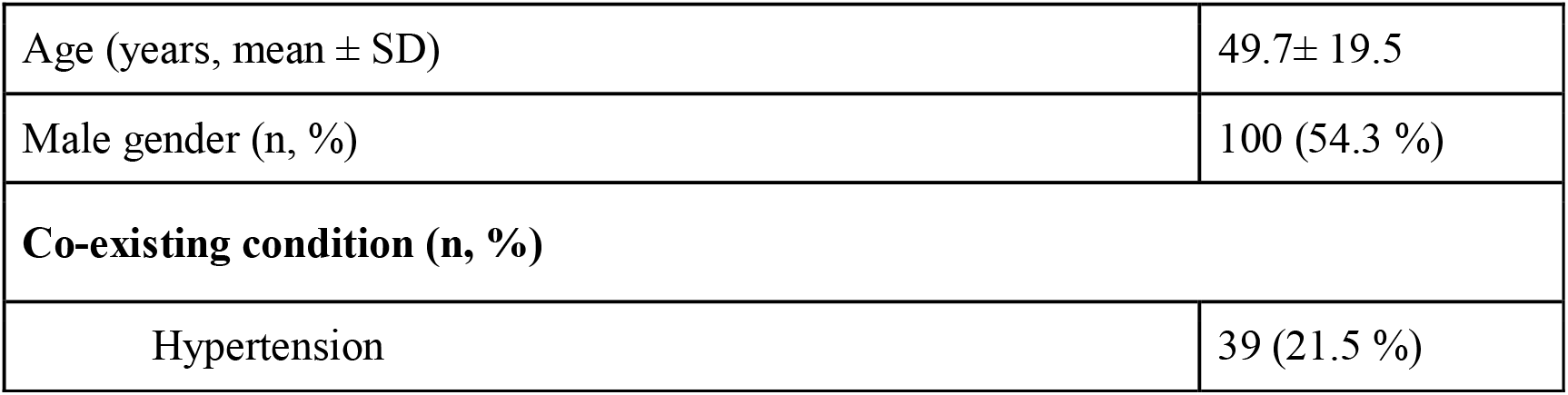
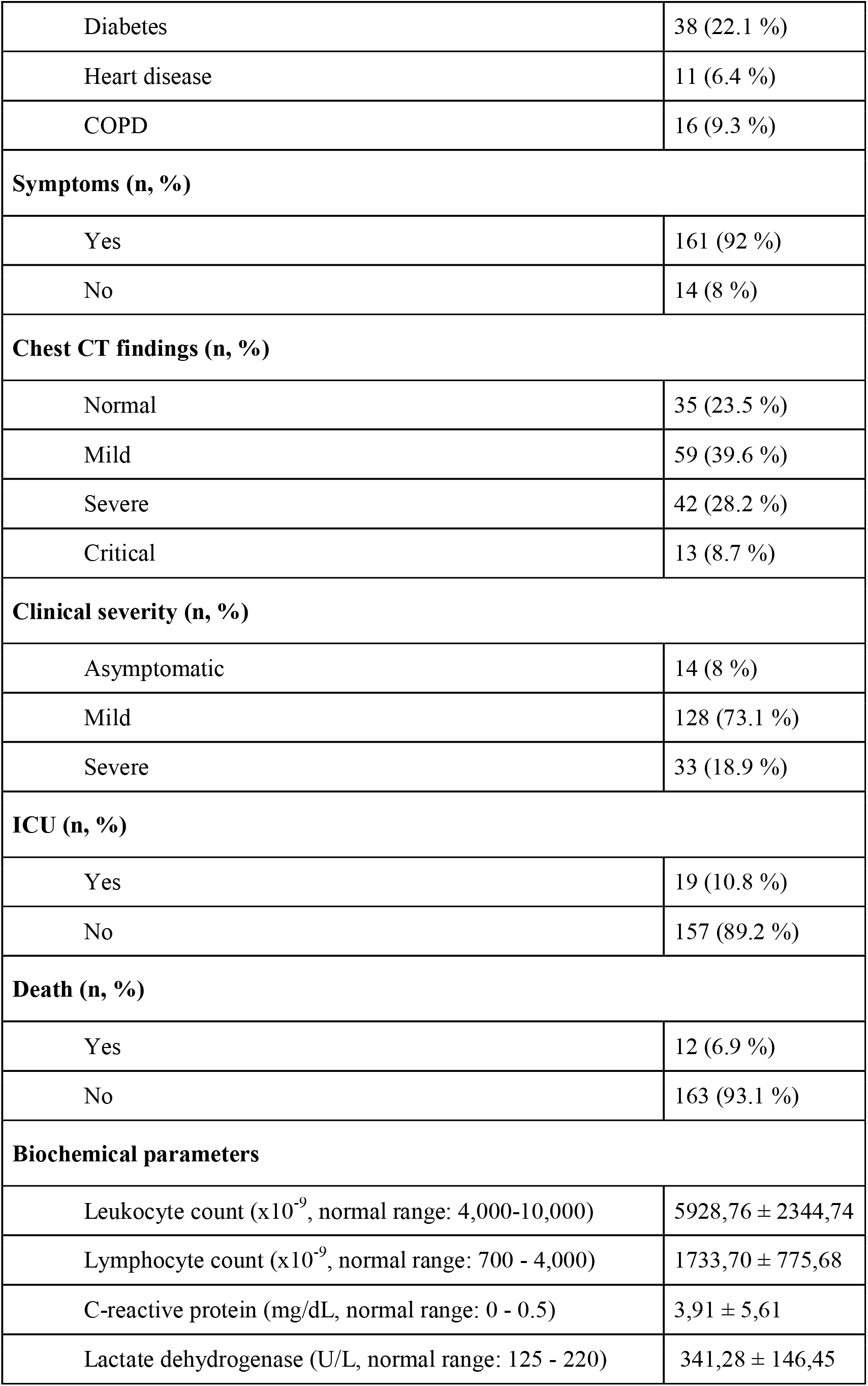
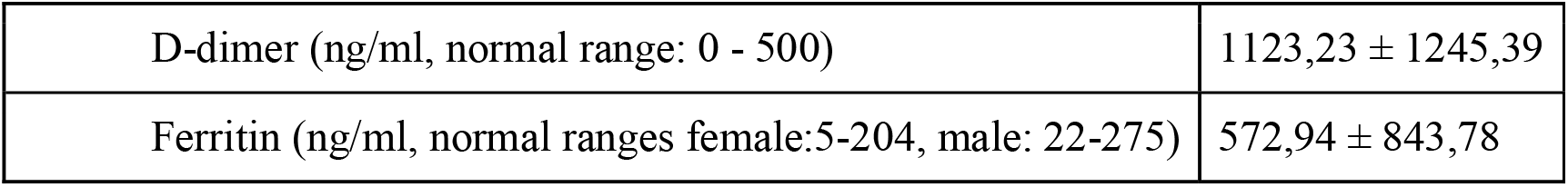
Demographic and clinical characteristics of 184 COVID-19 patients.

### Common viral variants and genetic phylogeny

Prepared amplicon libraries were sequenced on a NextSeq 500 instrument with an average output of 1.29±0.64 million reads per sample resulting in an average of 5,212 ± 3,931 depth of coverage. In total, 316 different nucleotide mutations were detected with an average count of 10.33 per sample. Among analyzed viral isolates, 175 were non-synonymous, 116 were synonymous, 16 were non-coding, six were insertion/deletion, and three were stop-gain mutations. The most predominant mutation was C>U transition which came up to 52.37% of total conversion. The most frequent mutation among 184 isolates and their consequences on the genome was given respectively in Table 2. Positions 241, 23403, 3037, and 14408 were found to harbor the highest mutation which reoccurred in almost 97% of viral isolates in this study. Clade assignment based on Nextstrain nomenclature showed that the most common clades were 20B (59.8%) and 20A (33.7%) whereas 19A (4.3%) and 20C (2.2%) were in the minority (Figure 1).

**Table 2:** The most common mutations and amino acid changes in SARS-CoV-2 genomes identified in this study.

| Position | Percentage in all samples | Change | Effect | Gene / Region | Amino acid change |
| --- | --- | --- | --- | --- | --- |
| 241 | 97.2 % | C /T | Non-coding | ORF1a | NA |
| 23403 | 97.2 % | A /G | missense | S | D/G |
| 3037 | 96.1 % | C /T | synonymous | ORF1a | F/F |
| 14408 | 95.6 % | C /T | missense | ORF1b | P/L |
| 28881-28882 | 61.4 % | GG/AA | missense | N | R/K |
| 28883 | 60.8 % | G /C | missense | N | G/R |
| 12809 | 30.9 % | C /T | missense | ORF1a | L/F |
| 25563 | 27.6 % | G/T | missense | ORF3a | Q/H |
| 18877 | 25.5 % | C/T | synonymous | ORF1b | L/L |
| 19839 | 23.3 % | T/C | synonymous | ORF1b | N/N |

**Figure 1:**
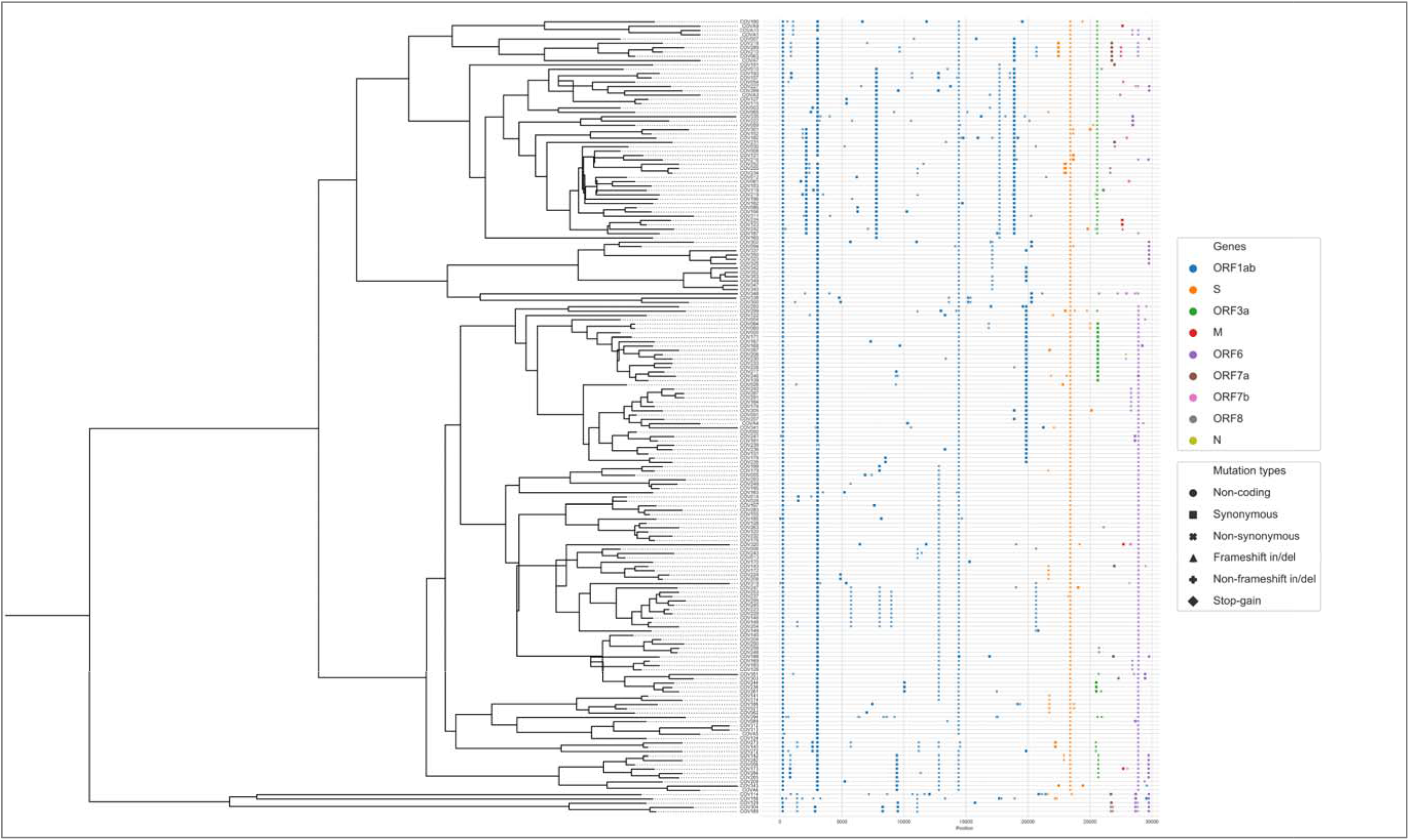
Phylogenetic tree of 184 viral genomes in this study together with mutations and their annotations.

### Viral clades and mutations are not associated with clinical outcome

Moreover, we compared the clinical findings of patients to the viral genome mutations and Nextstrain clades. We selected two clades (20A and 20B) due to obtaining enough sampling in our cohort and compared the clinical manifestations of the patients. As a result, patients infected by these two virus clades showed similar clinical features. Similarly, we found no statistical difference between viral genome mutations which were detected in at least 10 patients, and clinical severity, intensive care unit needs, and being asymptomatic. These results showed that commonly seen viral mutations and clades are not associated with clinical manifestations of patients in this study. Of note, the p.Q57H variant in the ORF3a gene has a significantly higher frequency in females (79.8%) compared to males (66%) with a p-value of 0.0472. Conversely, GGG to AAC trinucleotide change at positions 28881-28883 located in the N gene has been detected more frequently in males (48%) compared to females (30.9%) with a p-value of 0.0236. A complete list of tested variants and clinical parameters was given in Table 3.

**Table 3:**
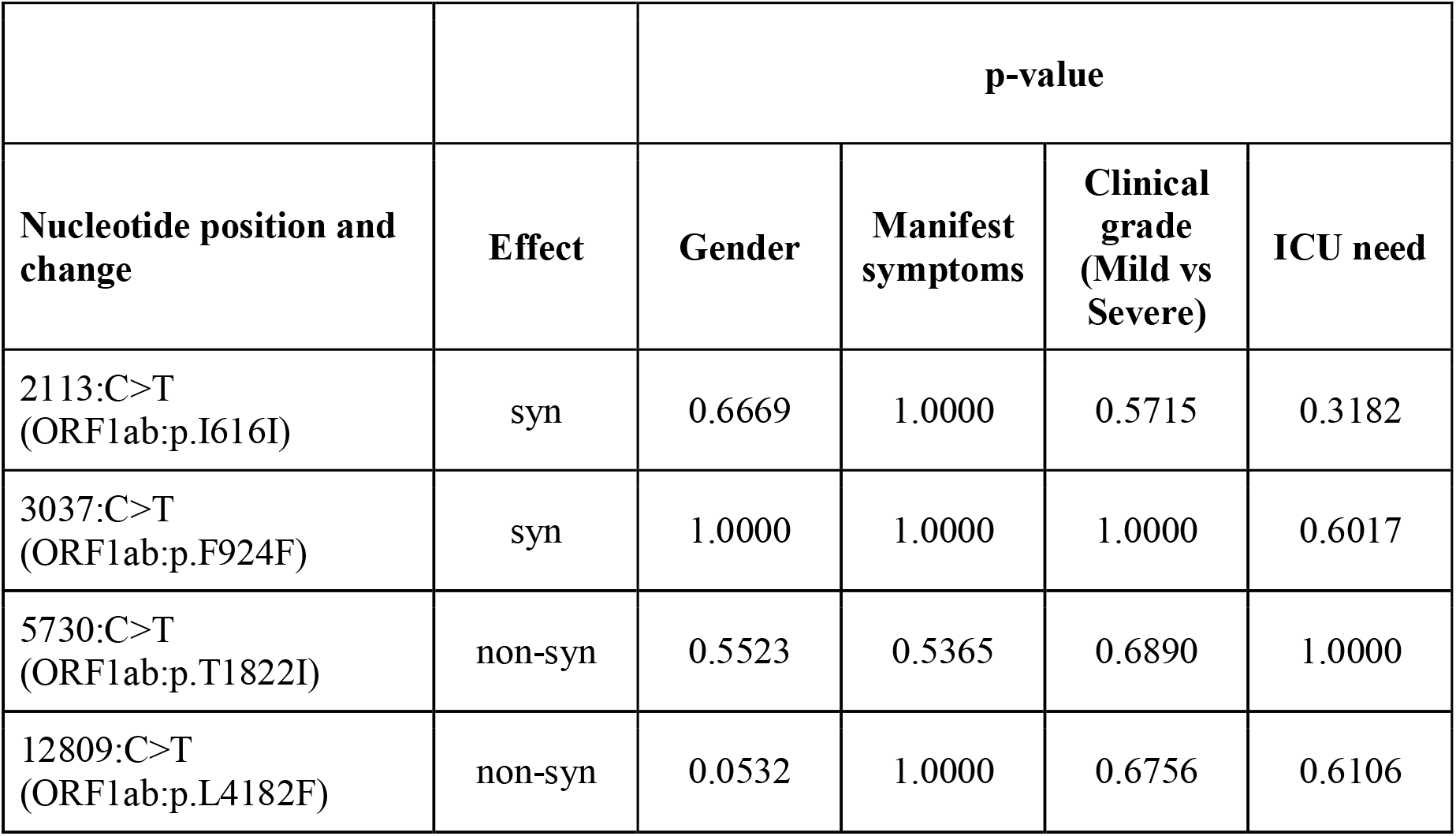
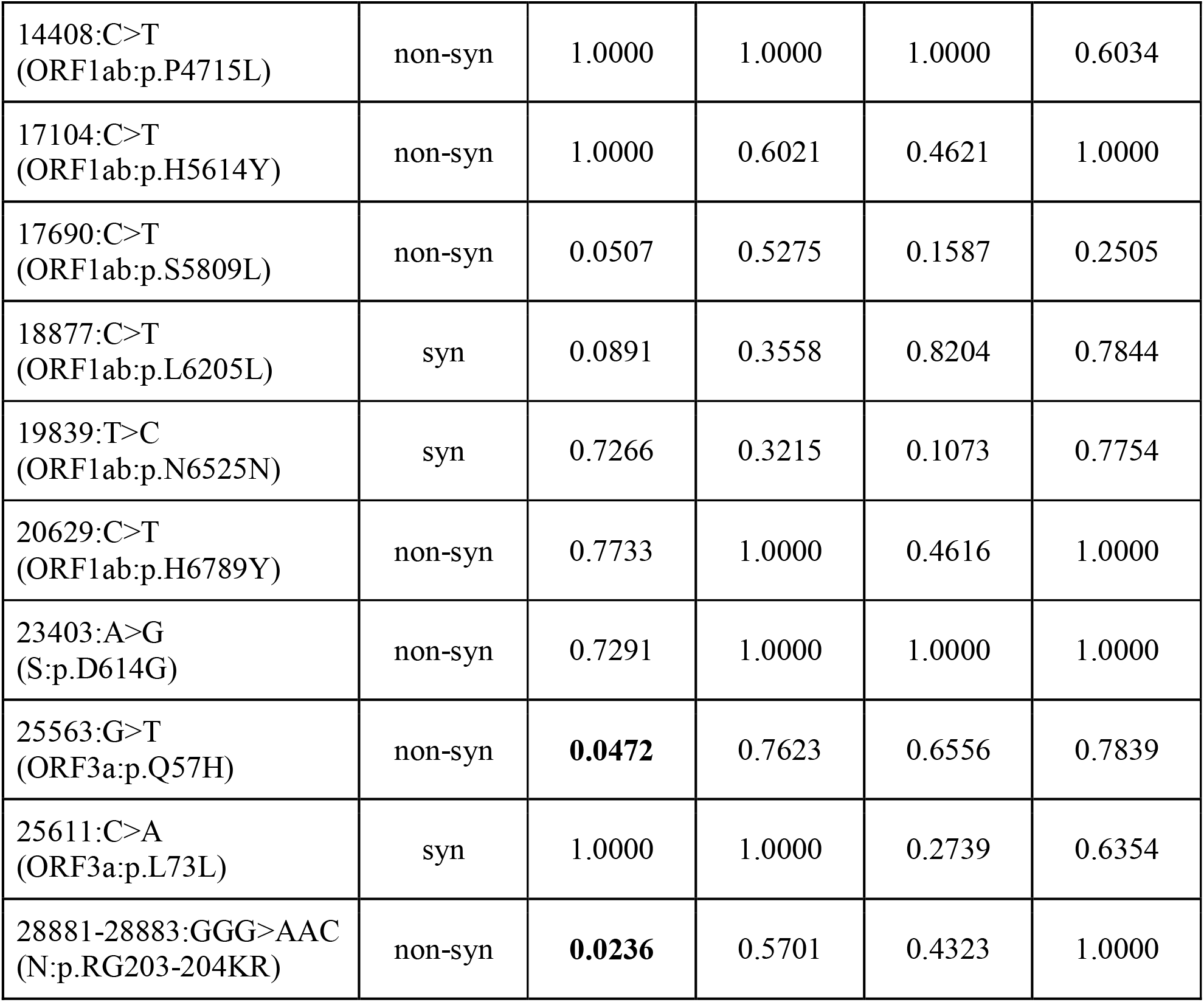
Selected SARS-CoV-2 variants in the study and two-sided Fisher’s exact test results.

### Possibly detrimental intra-host mutations are under negative selection

We generated consensus sequences of each isolate considering the major alleles (>50% of all reads) for genomic positions. Variant calling using NGS data revealed that 140 isolates (76%) have polymorphic sites in which minor allele frequencies (MAF) range between 10-90%. The rest of the samples have mutations only whose allelic frequency was over 90%. We further focused on those polymorphic sites for the presence of any association to clinical parameters. However, we could not detect any significant association between the number of polymorphic variations and clinical data. Overall, we identified 430 polymorphic sites in this study, and 95 (22.1%) of them were detected in at least two patients or identical to a consensus variant.

Further, we inspected those intra-host mutations for their occurrence events among isolates. Insertion and deletion mutations together with stop-gain mutations were more frequent in intra-host mutation events with statistical significance whereas non-coding variants were seen less in intra-host mutation events (Table 4). The overall frequencies of synonymous and non-synonymous mutation events were found not to be statistically significant between intra-host and consensus mutations. We also compared intra-host mutation counts to the clinical manifestations of the patients and no significant difference was detected.

**Table 4:** Amounts of intra-host and consensus mutation events among all 184 samples and their functional effects. AF: intra-host allelic frequency of the variant.

|  | Intra-host mutation events<br>(10% < AF < 90%) | Consensus mutations<br>(AF > 90%) | p-value |
| --- | --- | --- | --- |
| Non-synonymous | 469 | 857 | 1.0000 |
| Synonymous | 317 | 526 | 0.2621 |
| Insertion and deletion | 46 | 15 | **<0.0001** |
| Stop-gain | 11 | 2 | **0.0004** |
| Non-coding | 35 | 201 | **0.0001** |

## Discussion

SARS-CoV-2, which is an RNA virus, is spreading globally at an unprecedented speed. Naturally, it evolves and some mutations which may alter its virulence or infectivity can be expected. High throughput sequencing capacity of laboratories together with experiences with other pathogens give the research community the ability to follow-up this rapidly expanding infection closely. Genomics based viral outbreak monitoring gives chance to investigate transmission dynamics and clinical impact of mutations (Alm et al., 2020). Besides, chasing viral strains in the populations is very important not only observing local or global spread but also tracing any mutation that may worsen the clinical outcomes or creating the risk of vaccine escape in the future, too. The majority of the naturally occurring mutations are expected to be neutral but some may provide advantageous or deleterious to the pathogen. Deleterious mutations that result in decreased infectivity will quickly disappear; inversely advantageous mutations are possible to reach high frequency in the population. Of note, deciding whether a mutation is neutral or advantageous during a global pandemic can be sophisticated due to founder effect or sampling bias (Liu et al., 2020; Mavian et al., 2020; van Dorp et al., 2020). Herein, we analyzed whole-genome sequences of 184 SARS-CoV-2 isolates in Istanbul which was the epicenter of the pandemic in Turkey, using a high throughput sequencing method. We evaluated viral genomic changes for any association with the clinical outcomes of the patients.

A recent study pointed out that the majority of the analyzed samples in the European region consisted of 20A and 20B clades according to Nextstrain nomenclature (Alm et al., 2020). Similar to the European genomic surveillance study, 93.5% of samples analyzed in this work mostly belonged to the 20A or 20B clades. Isolates belonging to the 19A and 20C clades constituted 4.3% and 2.2% of all samples, respectively. Overall, we revealed that clade distribution in Istanbul was highly similar to that in Europe. Considering the pandemic outbreak in Turkey, it can be stated that the early introduction of the virus to Istanbul originated from Europe.

There have been criticisms of viral genetic differences and their effects on clinical outcomes of infected patients. Recent studies were unable to find any association between the clinical features of patients and viral mutations or clades. Even if there is a possibility for an arising mutation with potential pathological effects, there is not enough evidence to support this hypothesis, yet. Zhang et al. reported that major lineages among virus isolates have no major difference in clinical outcomes (Zhang et al., 2020). In this study, we are also unable to detect any genetic changes among sequenced viral genomes which may cause different clinical outcomes in terms of severity of the disease or manifesting symptoms. On the other hand, studies are suggesting viral genetic changes such as p.D614G mutation may play a role in pathogenicity or transmissibility (Hou et al., 2020; Korber et al., 2020). However, extensive genomic and functional studies are still needed to prove those findings. Together with some reports on the effects of p.D614G, the reason is still not known clearly whether this mutation became dominant due to evolutionary advantages, founder effect, or possible sampling bias in the western world (Grubaugh et al., 2020). Our dataset was underpowered to test any association between clinical manifestations and p.D614G mutation since more than 95% of our samples have this mutation.

Studies on SARS-CoV-2 genome variants are mainly focused to understand viral epidemiology, transmission dynamics, development of diagnostic assays, vaccines, and drugs. It is very difficult to determine the origins of each kind of genomic alteration, but large proportions of SARS-CoV-2 genome alterations are C>U changes (Matyasek and Kovarik, 2020; Simmonds, 2020). This type of nucleotide change was shown to be mainly caused by host RNA-editing mechanisms, namely ADAR and APOBEC deaminase mechanisms, as a part of the host antiviral defense mechanism (Di Giorgio et al., 2020). In our data, 52.39% of all conversions are C>U transitions. Since the deamination in humans is known to be a driver of mutational events and associated with a variety of diseases, clinical outcomes of COVID-19 patients might also be associated with this antiviral defense mechanism.

During the SARS-CoV-2 pandemic, high throughput sequencing has been globally used for viral genome analysis to uncover transmission patterns and evolutionary dynamics of the virus. Thus, this pandemic was the first in which researchers were able to continuously track genomic changes. Inter-host consensus sequences of the pathogen genome are always used for epidemiological investigations. However, these consensus sequences ignore the intra-host genomic variations of the virus population within the host. Besides all mutation events originated from an intra-host mutation, monitoring those events is also crucial to study its evolution dynamics. These intra-host variations of the virus genome may cause the immunological escape of the virus or affect the diagnostic performance of molecular assays. High-quality data of the deep sequencing methodologies give an opportunity to sensitively detect intra-host variations with minor allele frequencies (MAF) as low as 5%. In this study, we detected a total of 430 intra-host unique mutations with MAF between 10-90% in 76% of all samples. Our results indicated that subclonal mutations of the SARS-CoV-2 genome are prevalent among clinical samples. Also, 22.1% of those unique mutations occurred in at least two patients or identical to a consensus variant. This also indicates that the majority of those mutational events are not localized in hotspot sites.

Our results were highly similar to Rose et al. who reported intra-host mutations in 72% of analyzed 406 isolates (Rose et al., 2020). Of note, the number of intra-host variation events was not comparable since the conditions for the definition of polymorphic is not clear in the preprint article by Rose et al. whereas we used intra-host mutation definition if MAF was between 10% and 90% for a called variant.

A closer look at these polymorphic sites revealed that frequencies of non-synonymous and synonymous variants were not statistically different than inter-host variations (MAF > 90%) in the cohort. Whereas frequencies of stop-gain, in/del, and noncoding variants differ between intra- and inter-host variations. Stop-gain and in/del mutations were observed more in intra-host variations compared to inter-host consensus variants with MAF >90%. Insertion and deletions were detected at 5% and 0.9% in all intra- and inter-host variants, respectively. Similarly, the stop-gain variation rate in inter-host variations was 0.12% whereas its frequency in inter-host variations was ten times more: 1.2%. This result may be due to possible detrimental effects of those mutations by reducing viral activity.

Even though polymorphic sites might also be caused by amplification or sequencing errors, detection of those variants with different sequencing methodologies indicates the presence of intra-host variants. We also considered if these intra-host variations were caused by mixed infections but there is not any statistical difference in the occurrence of intra-host variations between outpatient and hospitalized groups. Further investigations about intra-host mutations are still required for understanding SARS-CoV-2 genome dynamics.

To conclude, our results were not able to identify any significant SARS-CoV-2 mutation related to the patient’s clinical manifestations or severity of the disease at this stage. Besides, we report mostly unique intra-host mutations in 76% of the samples with MAF ranging between 10-90 %, indicating continuous intra-host mutational events are very common. Since those mutations might be an origin for a lineage causing immunological escape of the virus, further longitudinal global genomic surveillance is needed.

## Data Availability

All sequence data of analyzed virus isolates were accessible through GISAID database.

## Acknowledgements

We thank the patients for their contribution to this study. This work was supported by a grant from the Health Institutes of Turkey (TUSEB Grant No: 8799).

## Author contributions

Conceptualization, L.D., G.D.D, I.K.; Data, T.K.A., N.B.A., M.A., N.D.C., A.S.O., N.A., A.I., Y.K.D., O.A.D.; Clinical sample management, T.K.A., N.B.A., P.Z., G.A., J.Y., B.K.; High-throughput sequencing, T.K.A., G.A., J.Y., B.K.; Analysis, I.K., P.Z.; Writing - Original Draft, I.K.; Writing - Review & Editing, I.K., N.B.A., L.D., G.D.D.

